# Effectiveness of Omicron XBB.1.5 vaccine against SARS-CoV-2 Omicron XBB and JN.1 infection in a prospective cohort study in the Netherlands, October 2023 to January 2024

**DOI:** 10.1101/2024.02.15.24302872

**Authors:** Anne J. Huiberts, Christina E. Hoeve, Brechje de Gier, Jeroen Cremer, Bas van der Veer, Hester E. de Melker, Janneke H.H.M. van de Wijgert, Susan van den Hof, Dirk Eggink, Mirjam J. Knol

## Abstract

We estimated vaccine effectiveness (VE) of SARS-CoV-2 Omicron XBB.1.5 vaccination against self-reported infection between 9 October 2023 and 9 January 2024 in 23,895 XBB.1.5 vaccine-eligible adults who had previously received at least one booster. VE was 41% (95%CI:23-55) in 18-59-year-olds and 50% (95%CI:44-56) in 60-85-year-olds. Sequencing data in a subset of infections suggests immune escape of the emerging BA.2.86 (JN.1) variant from recent prior infection (OR:2.6; 95%CI:1.1-6.3) and, although not statistically significant, from XBB.1.5 vaccination (OR:1.6; 95%CI:0.9-2.9).

A monovalent mRNA vaccine targeting the SARS-CoV-2 Omicron XBB.1.5 subvariant (Comirnaty) was used in the 2023 Dutch COVID-19 vaccination campaign that started on October 2, 2023. Individuals aged ≥60 years, medical risk groups, pregnant women and healthcare workers were eligible for vaccination. Since September 2023, a new Omicron BA.2.86 sub-variant named JN.1 has emerged and quickly became dominant in the Netherlands and globally [1, 2]. BA.2.86 is genetically divergent from the previously circulating XBB variants, indicating potential for immune escape [3].

We estimated vaccine effectiveness (VE) of XBB.1.5 vaccination against self-reported SARS-CoV-2 infection between 9 October 2023 and 9 January 2024 among adults aged 18-85 years who had previously received primary vaccination and at least one booster vaccination before 2 October 2023 and were eligible for XBB.1.5 vaccination. To assess potential immune escape by JN.1, we analyzed whether there was an association between XBB.1.5 vaccination or prior infection and the Omicron variant causing the infection (XBB vs. BA.2.86, including JN.1). The variant was determined by sequencing of viral genetic material present in positive lateral flow antigen self-tests.

## Study population

We used data from 23,895 participants of the VAccine Study COvid-19 (VASCO), an ongoing prospective cohort study among community-dwelling Dutch adults aged 18-85 years that started in May 2021 [4, 5]. Participants can report COVID-19 vaccinations and positive SARS-CoV-2 tests (PCR or lateral flow antigen (self-)test) any time. Participants provide self-collected fingerprick blood samples every 6 months for serological testing. Self-tests are provided to participants. For the current analysis, follow-up time started on 9 October 2023 and ended on either 9 January 2024, date of first positive SARS-CoV-2 test or date of last completed questionnaire, whichever occurred first. The seven person-days after XBB.1.5-vaccination were excluded. If participants reported a vaccination or infection in the three months prior to the XBB.1.5 vaccination campaign, follow-up started three months after this vaccination or infection.

Of 4,861 participants aged 18-59 years (XBB.1.5 vaccine-eligible because of being a healthcare worker or belonging to a medical risk group), 1,167 (24%) received XBB.1.5 vaccination during the follow-up period (**Table 1**). Of 19,034 participants aged 60-85 years (XBB.1.5 vaccine-eligible because of age), 11,330 participants (60%) received XBB.1.5 vaccination. In both age groups, XBB.1.5 vaccination recipients more often received a bivalent vaccination in the autumn 2022 COVID-19 vaccination campaign, compared to non-recipients (18-59-year-olds: 91% vs 56%; 60-85-year-olds: 93% vs 68%).

**Table 1.**
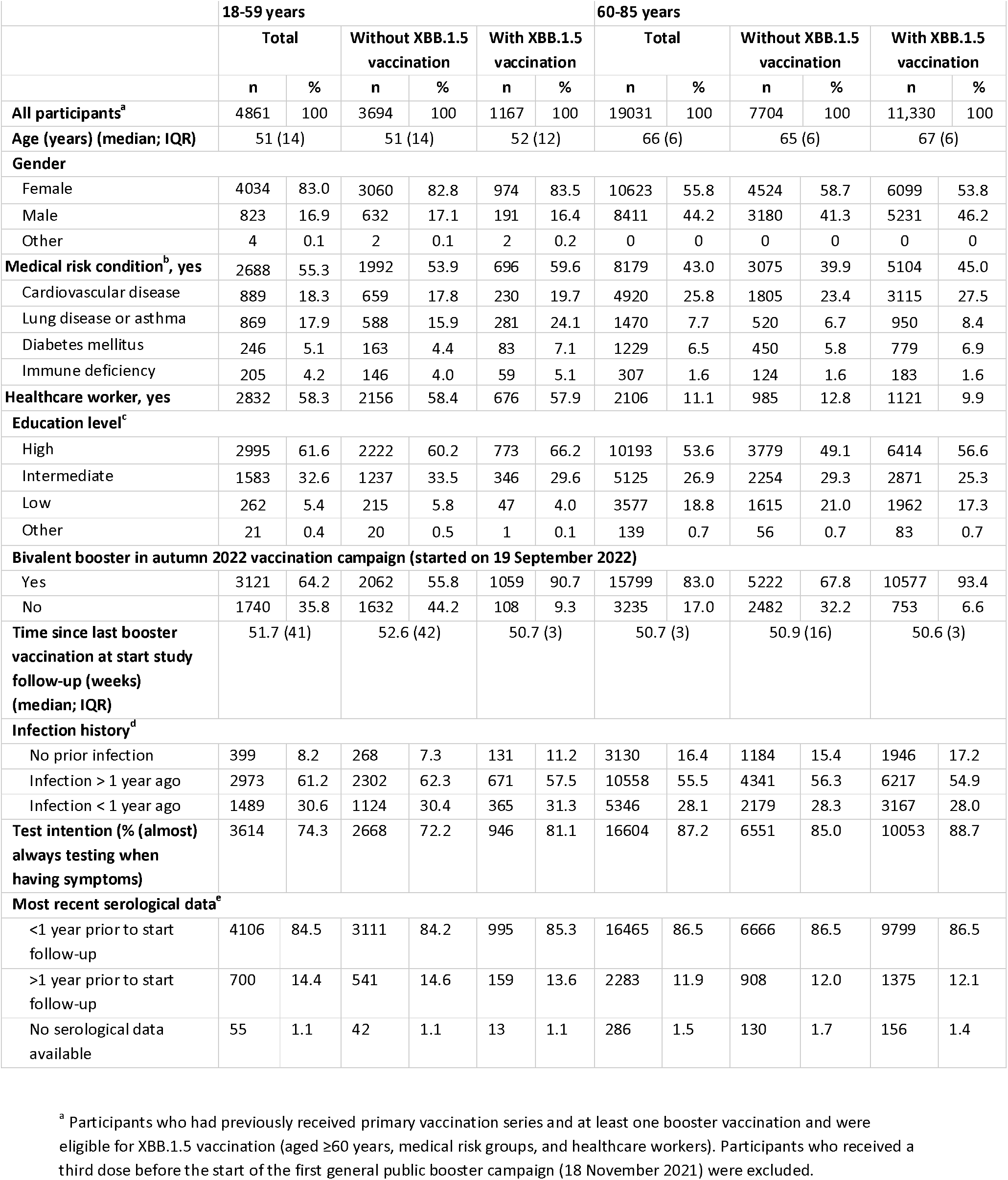
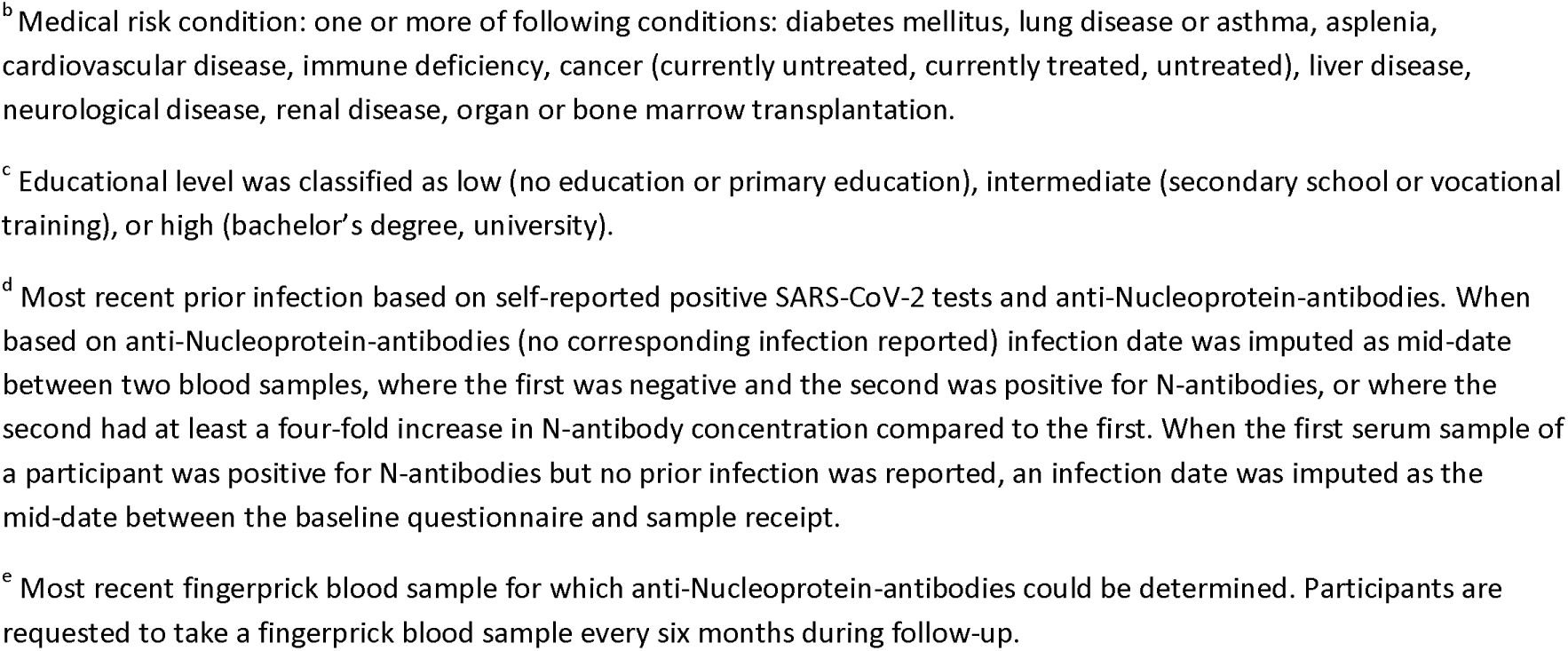
Characteristics of included VASCO participants vaccinated with primary series, at least one booster dose and eligible for XBB.1.5 vaccination.

|  | 18-59 years |  |  |  |  |  | 60-85 years |  |  |  |  |  |
| --- | --- | --- | --- | --- | --- | --- | --- | --- | --- | --- | --- | --- |
|  | Total |  | Without XBB.1.5 vaccination |  | With XBB.1.5 vaccination |  | Total |  | Without XBB.1.5 vaccination |  | With XBB.1.5 vaccination |  |
|  | n | % | n | % | n | % | n | % | n | % | n | % |
| All participants<sup>a</sup> | 4861 | 100 | 3694 | 100 | 1167 | 100 | 19031 | 100 | 7704 | 100 | 11,330 | 100 |
| Age (years) (median; IQR) | 51 (14) |  | 51 (14) |  | 52 (12) |  | 66 (6) |  | 65 (6) |  | 67 (6) |  |
| Gender |  |  |  |  |  |  |  |  |  |  |  |  |
| Female | 4034 | 83.0 | 3060 | 82.8 | 974 | 83.5 | 10623 | 55.8 | 4524 | 58.7 | 6099 | 53.8 |
| Male | 823 | 16.9 | 632 | 17.1 | 191 | 16.4 | 8411 | 44.2 | 3180 | 41.3 | 5231 | 46.2 |
| Other | 4 | 0.1 | 2 | 0.1 | 2 | 0.2 | 0 | 0 | 0 | 0 | 0 | 0 |
| Medical risk condition<sup>b</sup>, yes | 2688 | 55.3 | 1992 | 53.9 | 696 | 59.6 | 8179 | 43.0 | 3075 | 39.9 | 5104 | 45.0 |
| Cardiovascular disease | 889 | 18.3 | 659 | 17.8 | 230 | 19.7 | 4920 | 25.8 | 1805 | 23.4 | 3115 | 27.5 |
| Lung disease or asthma | 869 | 17.9 | 588 | 15.9 | 281 | 24.1 | 1470 | 7.7 | 520 | 6.7 | 950 | 8.4 |
| Diabetes mellitus | 246 | 5.1 | 163 | 4.4 | 83 | 7.1 | 1229 | 6.5 | 450 | 5.8 | 779 | 6.9 |
| Immune deficiency | 205 | 4.2 | 146 | 4.0 | 59 | 5.1 | 307 | 1.6 | 124 | 1.6 | 183 | 1.6 |
| Healthcare worker, yes | 2832 | 58.3 | 2156 | 58.4 | 676 | 57.9 | 2106 | 11.1 | 985 | 12.8 | 1121 | 9.9 |
| Education level<sup>c</sup> |  |  |  |  |  |  |  |  |  |  |  |  |
| High | 2995 | 61.6 | 2222 | 60.2 | 773 | 66.2 | 10193 | 53.6 | 3779 | 49.1 | 6414 | 56.6 |
| Intermediate | 1583 | 32.6 | 1237 | 33.5 | 346 | 29.6 | 5125 | 26.9 | 2254 | 29.3 | 2871 | 25.3 |
| Low | 262 | 5.4 | 215 | 5.8 | 47 | 4.0 | 3577 | 18.8 | 1615 | 21.0 | 1962 | 17.3 |
| Other | 21 | 0.4 | 20 | 0.5 | 1 | 0.1 | 139 | 0.7 | 56 | 0.7 | 83 | 0.7 |
| Bivalent booster in autumn 2022 vaccination campaign (started on 19 September 2022) |  |  |  |  |  |  |  |  |  |  |  |  |
| Yes | 3121 | 64.2 | 2062 | 55.8 | 1059 | 90.7 | 15799 | 83.0 | 5222 | 67.8 | 10577 | 93.4 |
| No | 1740 | 35.8 | 1632 | 44.2 | 108 | 9.3 | 3235 | 17.0 | 2482 | 32.2 | 753 | 6.6 |
| Time since last booster vaccination at start study follow-up (weeks) (median; IQR) | 51.7 (41) |  | 52.6 (42) |  | 50.7 (3) |  | 50.7 (3) |  | 50.9 (16) |  | 50.6 (3) |  |
| Infection history<sup>d</sup> |  |  |  |  |  |  |  |  |  |  |  |  |
| No prior infection | 399 | 8.2 | 268 | 7.3 | 131 | 11.2 | 3130 | 16.4 | 1184 | 15.4 | 1946 | 17.2 |
| Infection > 1 year ago | 2973 | 61.2 | 2302 | 62.3 | 671 | 57.5 | 10558 | 55.5 | 4341 | 56.3 | 6217 | 54.9 |
| Infection < 1 year ago | 1489 | 30.6 | 1124 | 30.4 | 365 | 31.3 | 5346 | 28.1 | 2179 | 28.3 | 3167 | 28.0 |
| Test intention (% (almost) always testing when having symptoms) | 3614 | 74.3 | 2668 | 72.2 | 946 | 81.1 | 16604 | 87.2 | 6551 | 85.0 | 10053 | 88.7 |
| Most recent serological data<sup>e</sup> |  |  |  |  |  |  |  |  |  |  |  |  |
| <1 year prior to start follow-up | 4106 | 84.5 | 3111 | 84.2 | 995 | 85.3 | 16465 | 86.5 | 6666 | 86.5 | 9799 | 86.5 |
| >1 year prior to start follow-up | 700 | 14.4 | 541 | 14.6 | 159 | 13.6 | 2283 | 11.9 | 908 | 12.0 | 1375 | 12.1 |
| No serological data available | 55 | 1.1 | 42 | 1.1 | 13 | 1.1 | 286 | 1.5 | 130 | 1.7 | 156 | 1.4 |

<sup>a</sup> Participants who had previously received primary vaccination series and at least one booster vaccination and were eligible for XBB.1.5 vaccination (aged ≥60 years, medical risk groups, and healthcare workers). Participants who received a third dose before the start of the first general public booster campaign (18 November 2021) were excluded.
<sup>b</sup> Medical risk condition: one or more of following conditions: diabetes mellitus, lung disease or asthma, asplenia, cardiovascular disease, immune deficiency, cancer (currently untreated, currently treated, untreated), liver disease, neurological disease, renal disease, organ or bone marrow transplantation.
<sup>c</sup> Educational level was classified as low (no education or primary education), intermediate (secondary school or vocational training), or high (bachelor's degree, university).
<sup>d</sup> Most recent prior infection based on self-reported positive SARS-CoV-2 tests and anti-Nucleoprotein-antibodies. When based on anti-Nucleoprotein-antibodies (no corresponding infection reported) infection date was imputed as mid-date between two blood samples, where the first was negative and the second was positive for N-antibodies, or where the second had at least a four-fold increase in N-antibody concentration compared to the first. When the first serum sample of a participant was positive for N-antibodies but no prior infection was reported, an infection date was imputed as the mid-date between the baseline questionnaire and sample receipt.
<sup>e</sup> Most recent fingerprick blood sample for which anti-Nucleoprotein-antibodies could be determined. Participants are requested to take a fingerprick blood sample every six months during follow-up.

During follow-up, 1,951 SARS-CoV-2 infections were reported. Reported incidence increased during the study period (**Figure 1**), consistent with national syndromic and wastewater surveillance data [6, 7]. Those without prior infection, defined as no self-reported positive SARS-CoV-2 test and negative anti-Nucleoprotein serology (8% of 18-59-year-olds; 16% of 60-85-year-olds), had the highest incidence (note that 18-59-year-olds included low number of participants and infections; **Figure S1**). Incidence was lowest among those with a prior infection <1 year ago. Incidence was consistently lower among those with XBB.1.5 vaccination compared to those without regardless of prior infection history.

**Figure 1.**
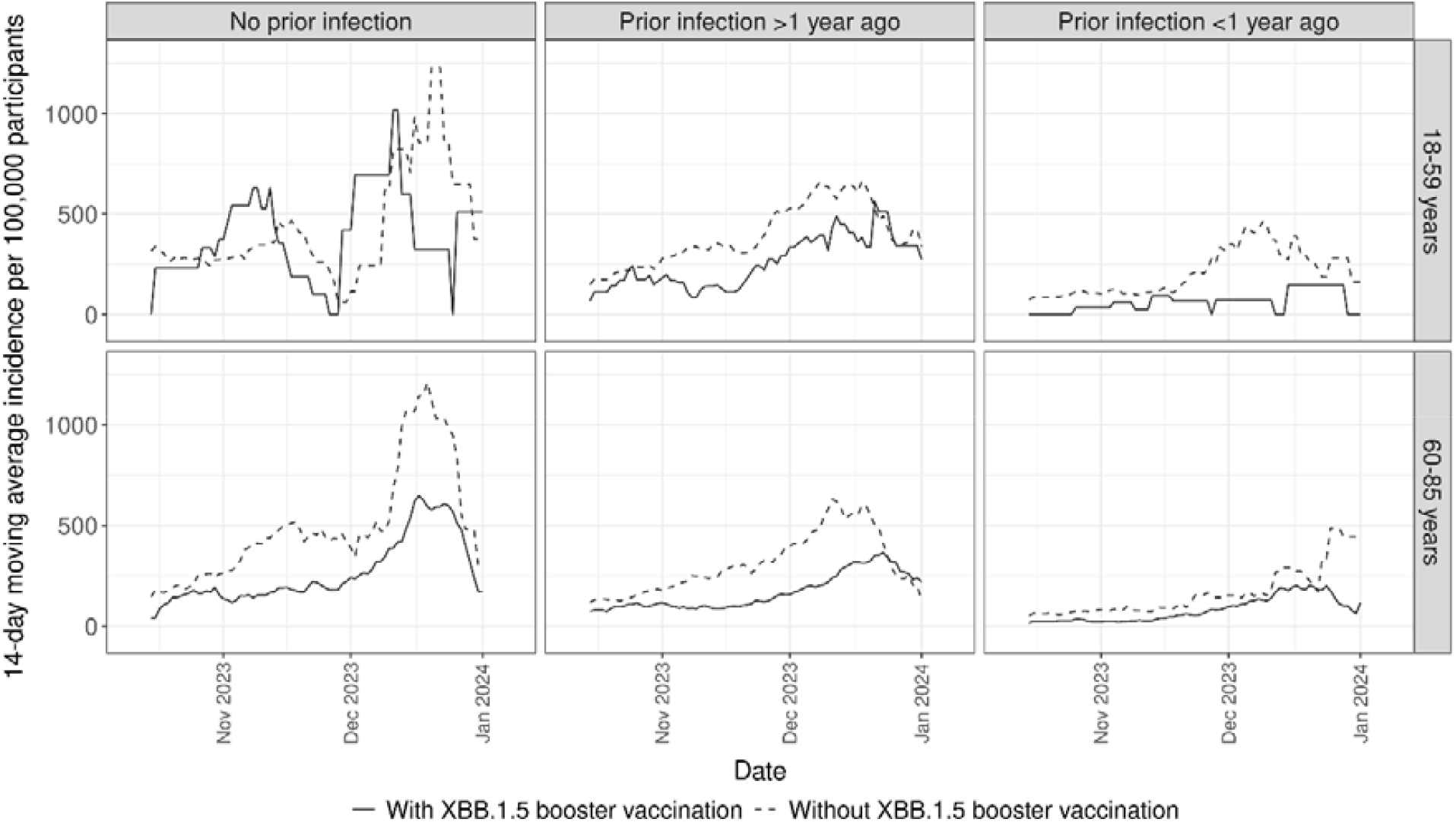
14-day moving average of the number of reported infections per 100,000 participants in the period 9 October 2023 to 9 January 2024, by infection history and age group

### Vaccine effectiveness

VE was estimated using Cox proportional hazard models with calendar time as time scale, XBB.1.5-vaccination as time-varying exposure and adjustment for age group, sex, education level, medical risk condition and infection history. Analyses were performed using R version 4.3.2, packages Epi, survival and stats.

VE was 41% (95%CI:23-55) among 18-59-year-olds and 50% (95%CI:44-56) among 60-85-year-olds (**Figure 2, Table S1**). VE estimates were somewhat higher up to 6 weeks after vaccination compared to 7-12 weeks after vaccination among 60-85 year-olds (52% vs 41%), but CIs are wide. A sensitivity analysis among participants who, during the study period, reported to (almost) always test in case of symptoms showed a slightly higher VE of 45% (95%CI:26-59) among 18-59-year-olds and 54% (95%CI:47-59) among 60-85-year-olds (**Figure 2, Table S2**). When only known symptomatic infections (68% of the infections; 21% asymptomatic, 11% unknown) were included, VE was 35% among 18-59-year-olds and 55% among 60-85-year-olds.

**Figure 2.**
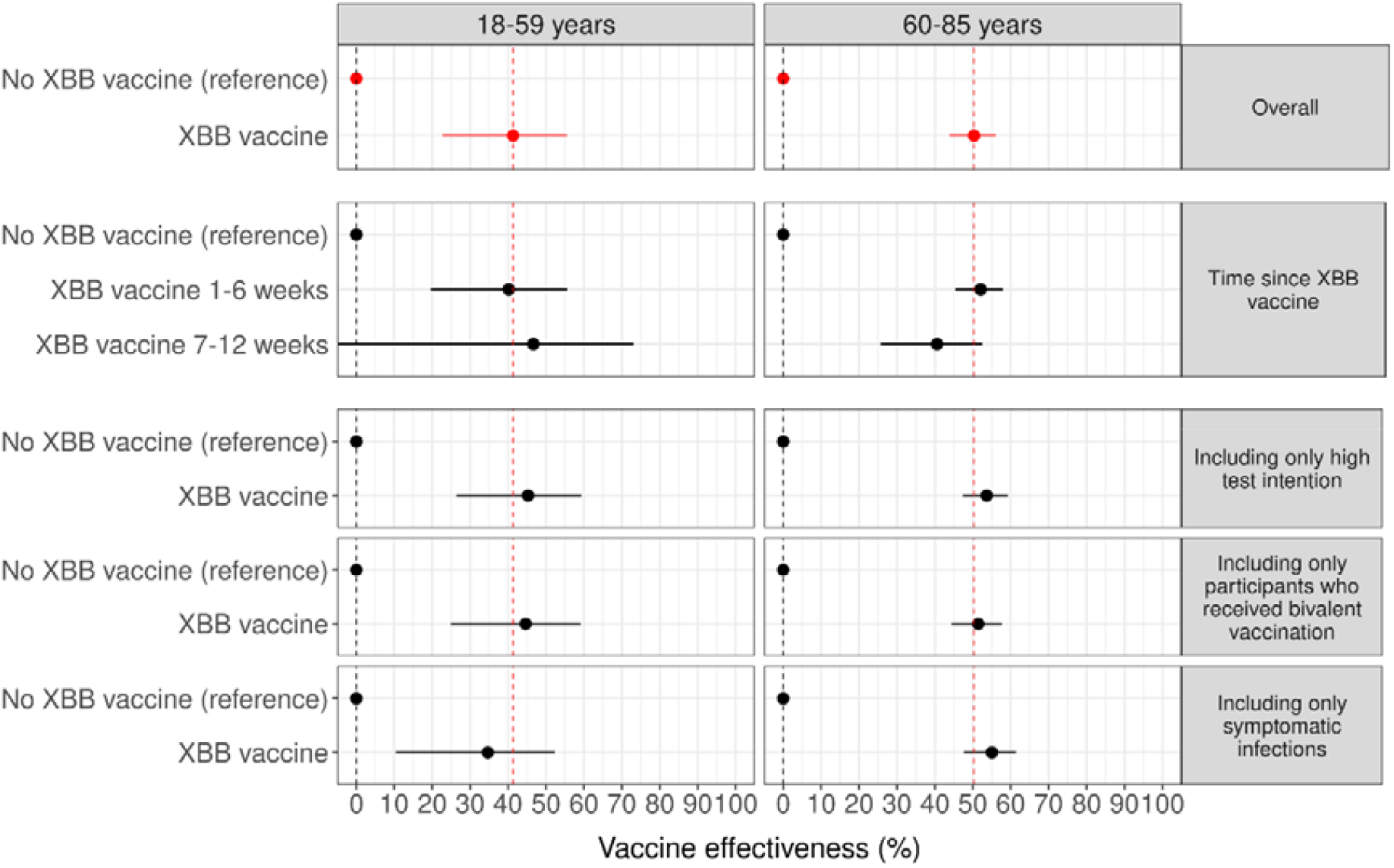
Vaccine effectiveness^a^ and 95% confidence interval of XBB.1.5 vaccine against Omicron infection in the period 9 October 2023 to 9 January 2024 among participants vaccinated with primary series and at least one booster and eligible for XBB.1.5 vaccination stratified by age group ^a^Adjusted for age group (18-39, 40-59, 60-69, 70-85 years), sex, education level, presence of a medical risk condition and infection history

Independent of infection history, XBB.1.5 vaccination provided additional protection in both 18-59-year-olds and 60-85-year-olds. Among 60-85-year-olds, protection against infection was highest after a prior infection <1 year ago either with (85%) or without (73%) XBB.1.5 vaccination (**Table S2**). This same trend was seen in 18-59-year-olds (87% and 61%, respectively).

### Emerging subvariant

From June 2023, VASCO participants were asked to submit positive self-tests for sequencing. Residual viral genetic material was extracted from the test strips of the self-tests (as described by [8]), and sequenced if the RT-PCR reactions were positive, to determine the variant that caused the infection [9].

Of 3,525 reported infections between 9 October 2023 and 9 January 2024 (also including infections notified after completion of the last questionnaire and therefore excluded from the VE analysis), 764 self-tests with a strong positive band were sequenced (**Figure S2**). Of those, 530 resulted in (near) whole genome sequences. Identified variants were Omicron XBB (149;28%), Omicron BA.2.86 (370;70%), of which 314 (85%) were sub-variant JN.1, and other Omicron variants (11;2%). Infections caused by BA.2.86 increased from 13% in the first week of the study period to 100% in the last week (**Figure S3**). Observed prevalence of variants corresponded well with national SARS-CoV-2 genomic surveillance [10].

Among infected participants with sequencing data (**Table S3**), we performed logistic regression to estimate the association between XBB.1.5 vaccination or prior infection and the Omicron variant causing the infection (XBB and BA.2.86), adjusting for testing week, age group, sex, education level and medical risk condition. By adjusting for testing week, we compared XBB and BA.2.86 infections at the same moment in calendar time.

XBB.1.5 vaccinated participants had a non-significantly higher odds of their infection being caused by BA.2.86 rather than XBB (OR:1.6; 95%CI:0.9-2.9); **Table 2**), suggesting that the VE against BA.2.86 may be lower than the VE against XBB. Participants with a prior infection <1 year ago had a significantly higher odds of their infection being caused by BA.2.86 rather than XBB (OR:2.6; 95%CI:1.1-6.3), compared to participants without prior infection. No significant association was observed for participants with a prior infection >1 year ago (OR:1.4; 95%CI:0.7-2.5).

**Table 2.** Association between XBB.1.5 vaccination status and SARS-CoV-2 BA.2.86 vs XBB infection, in the period 9 October 2023 to 9 January 2024.

|  | XBB infection |  | BA.2.86 infection |  | Adjusted<sup>a</sup> OR (95% CI) |
| --- | --- | --- | --- | --- | --- |
|  | n | % | n | % |  |
| **XBB.1.5 vaccination status** |  |  |  |  |  |
| Without XBB.1.5 vaccination | 118 | 79.2 | 173 | 46.8 | Reference |
| With XBB.1.5 vaccination | 31 | 20.8 | 197 | 53.2 | 1.6 (0.9-2.9) |
| **Infection history** |  |  |  |  |  |
| No prior infection | 35 | 23.5 | 65 | 17.6 | Reference |
| Infection >1 year ago | 98 | 65.8 | 229 | 61.9 | 1.4 (0.7-2.5) |
| Infection <1 year ago | 16 | 10.7 | 76 | 20.5 | 2.6 (1.1-6.3) |

<sup>a</sup> Adjusted for testing week, age group (18-39, 40-59, 60-69, 70-85), sex, education level and presence of a medical risk condition

## Discussion and conclusion

The VE of XBB.1.5 vaccination against self-reported SARS-CoV-2 infection among XBB.1.5 vaccine-eligible participants who had previously received a primary vaccination series and at least one booster vaccination was 41% for 18-59-year-olds and 50% for 60-85-year-olds, with similar estimates against symptomatic infection. Furthermore, irrespective of vaccination status, protection of a recent prior infection against a new infection was high. Protection from XBB.1.5 vaccination and recent prior infection seemed lower against BA.2.86 compared to XBB infection (OR of 1.6 (non-significant) and 2.6, respectively), suggesting immune escape by BA.2.86.

A recent study from the US reported comparable estimates against symptomatic infection of 57% in 18-49-year-olds and 46% in those aged ≥50 years [11]. Their VE estimates were considerably higher compared to their estimates during the 2022/2023 winter season of the bivalent booster vaccination (46%, 38% and 36% in individuals aged 18–49 years, 50–64 and ≥ 65 years, respectively) [12]. Also our estimates were considerably higher compared to our estimates of the bivalent booster: 31% in 18-59-year-olds and 14% in 60-85-year-olds [13]. This suggests that the 2023 XBB.1.5 vaccine better matches the virus that is circulating in the 2023/2024 winter season than the 2022 bivalent vaccine did in the 2022/2023 winter season, when Omicron BQ.1 and BA.2.75 were circulating in the Netherlands [10]. Possible alternative explanations for the higher VE compared to last year are the longer time period since prior vaccination campaigns and lower virus circulation in the summer of 2023. However, sensitivity analyses showed similar VEs among those who had received the bivalent vaccination, and after stratifying by infection history.

Preliminary immunological data suggest that BA.2.86 and JN.1 show modest signs of immune evasion to XBB.1.5 vaccination, but both strains were found to be neutralized by XBB.1.5 vaccine-induced antibodies [14, 15] and prior XBB infection [16]. A recent study from Denmark reported that XBB.1.5 vaccinated cases had a 1.6 times higher odds of their infection being caused by BA.2.86 than other variants compared to unvaccinated cases [17], which is consistent with our findings. Also, VE estimates of XBB.1.5 vaccination from the US were reported to be lower against infections caused by JN.1 (49%) than infections caused by XBB-related lineages (60%), although the difference was non-significant [11]. The apparent immunological advantage of the BA.2.86 sub-variant JN.1 may be the reason that it became dominant quickly, in contrast to different XBB subvariants that circulated simultaneously last year.

We provided participants with self-tests free-of-charge, thereby reducing bias associated with access to testing. However, we did rely on participant adherence to study instructions related to testing and reporting of infections, which could have influenced our estimates. However, restricting the analysis to participants with high test intention only slightly increased our estimates. Furthermore, serological data enabled us to detect and adjust for prior untested (asymptomatic) infections, but serology data were not available for all study participants (84-87% of participants in the past year). Furthermore, imputation of the infection date when an infection was detected by serology only may have resulted in misclassification of time since prior infection. Self-reported vaccination data was confirmed by linkage to vaccination registry data, limiting exposure misclassification.

Other studies have shown benefit of the XBB.1.5 vaccination campaign in reducing COVID-19 hospitalizations [18, 19]. We showed that XBB.1.5 vaccination also provided considerable protection against SARS-CoV-2 infection in the first three months post-vaccination. Recent prior infection also protects against new infection, but it should be kept in mind that experiencing a SARS-CoV-2 infection carries risk of severe disease in vulnerable groups, and of post-COVID condition. We found indications of immune escape of the emerging BA.2.86 (JN.1) variant from XBB.1.5 vaccination and prior infection, possibly explaining the rapid increase of this variant worldwide.

## Data Availability

Anonymized data reported from this study can be obtained from the corresponding author upon request. The dataset may include individual data and a data dictionary will be provided. Data requests should include a proposal for the planned analyses. Data transfer will require a signed data sharing agreement.

