## Supplementary file for "Effectiveness of Omicron XBB.1.5 vaccine against SARS-CoV-2 Omicron XBB and JN.1 infection in a prospective cohort study in the Netherlands, October 2023 to January 2024"

**Figure S1**. Number of participants and reported SARS-CoV-2 infections in the period 9 October 2023 to 9 January 2024, by vaccination status, infection history and age group

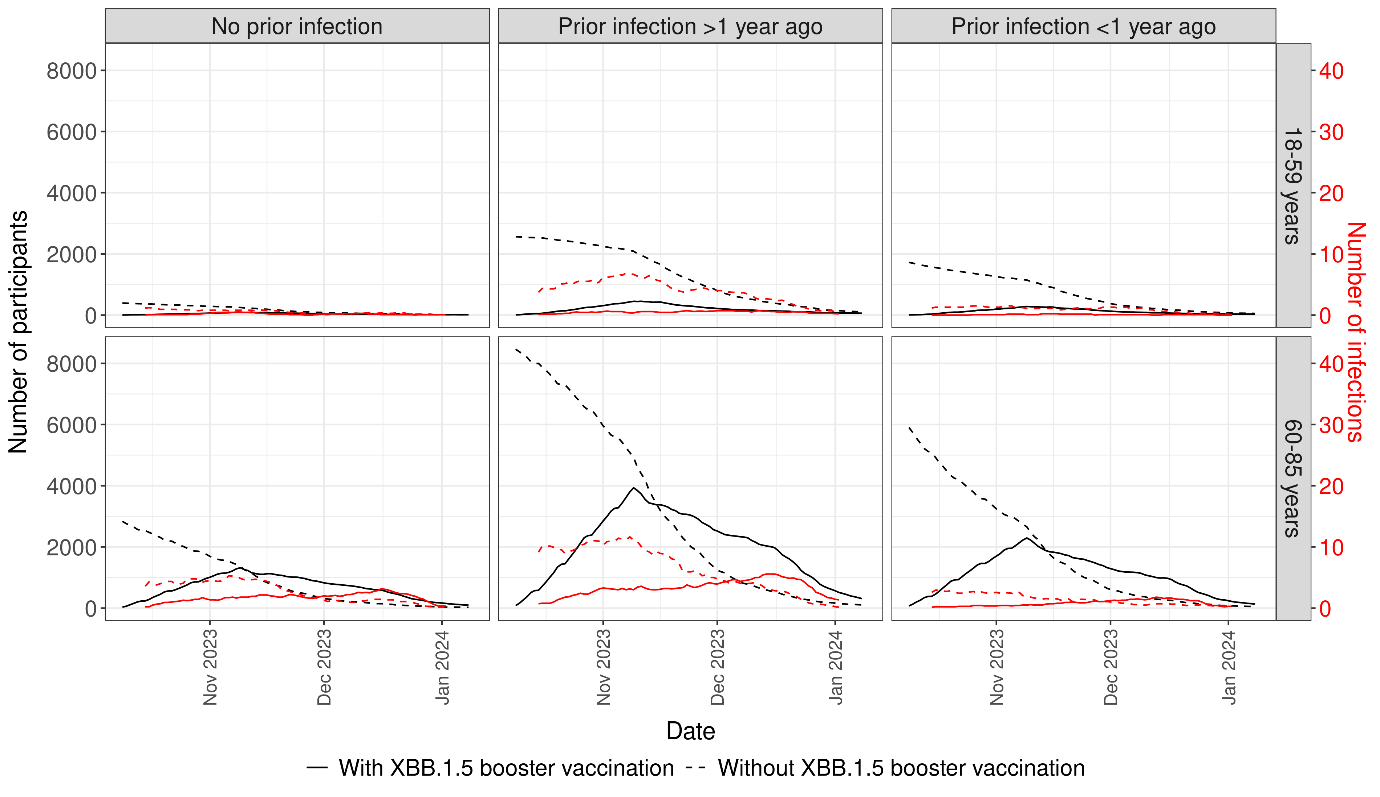

**Table S1**. Incidence rate and vaccine effectiveness of XBB.1.5 vaccination in the period 9 October 2023 to 9 January 2024, stratified by age group

|  | **Person-weeks** | **Number of infections** | **Rate (per 1,000 weeks)** | **Adjusted^a^ vaccine effectiveness (%) (95% CI)** |
| --- | --- | --- | --- | --- |
| **18-59 years** | | | | |
| **Overall^b^** | | | | |
| Without XBB vaccination | 28667 | 475 | 16.6 | Reference |
| With XBB vaccination | 4705 | 59 | 12.5 | 41.3 (22.6-55.5) |
| **By time since XBB vaccination** | | | | |
| Without XBB vaccination | 28667 | 475 | 16.6 | Reference |
| With XBB vaccination (1-6 weeks) | 4145 | 50 | 12.1 | 40.2 (19.6-55.5) |
| With XBB vaccination (7-12 weeks) | 552 | 9 | 16.3 | 46.7 (-5.7-73.1) |
| **Participants who (almost) always test in case of symptoms** | | | | |
| Without XBB vaccination | 20223 | 397 | 19.6 | Reference |
| With XBB vaccination | 3704 | 52 | 14.0 | 45.3 (26.4-59.3) |
| **Participants who received a bivalent booster in autumn 2022 COVID-19 vaccination campaign (started on 19 September 2022)** | | | | |
| Without XBB vaccination | 16492 | 284 | 17.2 | Reference |
| With XBB vaccination | 4274 | 53 | 12.4 | 44.6 (25.0-59.1) |
| **Symptomatic infections only** | | | | |
| Without XBB vaccination | 28667 | 330 | 11.5 | Reference |
| With XBB vaccination | 4705 | 46 | 9.8 | 34.7 (10.4-52.4) |
| **60-85 years** | | | | |
| **Overall** | | | | |
| Without XBB vaccination | 78581 | 939 | 11.9 | Reference |
| With XBB vaccination | 49512 | 478 | 9.7 | 50.3 (43.8-56.1) |
| **By time since XBB vaccination** | | | | |
| Without XBB vaccination | 78581 | 939 | 11.9 | Reference |
| With XBB vaccination (1-6 weeks) | 42819 | 356 | 8.3 | 52.1 (45.4-57.9) |
| With XBB vaccination (7-12 weeks) | 6666 | 122 | 18.3 | 40.6 (25.7-52.4) |
| **Participants who (almost) always test in case of symptoms** | | | | |
| Without XBB vaccination | 66839 | 878 | 13.1 | Reference |
| With XBB vaccination | 43742 | 448 | 10.2 | 53.7 (47.3-59.2) |
| **Participants who received a bivalent booster in autumn 2022 COVID-19 vaccination campaign (started on 19 September 2022)** | | | | |
| Without XBB vaccination | 58948 | 706 | 12.0 | Reference |
| With XBB vaccination | 46591 | 458 | 9.8 | 51.4 (44.3-57.6) |
| **Symptomatic infections only** | | | | |
| Without XBB vaccination | 78581 | 648 | 8.2 | Reference |
| With XBB vaccination | 49512 | 298 | 6.0 | 55.0 (47.6-61.4) |

^a^ Adjusted for age group (18-39, 40-59, 60-69, 70-85), sex, education level, medical risk condition and infection history.

^b^ Interaction between XBB.1.5 vaccination and infection history was not significant (p=0.17 and p=0.77 for 18-59 and 60-85 year-olds, respectively); therefore results were not stratified by infection history

**Table S2**. Protection^a^ by XBB.1.5 vaccination and prior infection in the period 9 October 2023 to 9 January 2024, stratified by age group

|  | **No prior infection** | **Prior infection >1 year ago** | **Prior infection <1 year ago** |
| --- | --- | --- | --- |
| **18-59 years** |  |  |  |
| Without XBB.1.5 vaccination | Reference | 14.0 (-16.5-36.6) | 61.3 (44.8-72.9) |
| With XBB.1.5 vaccination | 11.7 (-60.9-51.6) | 49.7 (22.8-67.2) | 86.7 (68.9-94.3) |
| **60-85 years** |  |  |  |
| Without XBB.1.5 vaccination | Reference | 32.9 (21.8-42.3) | 72.6 (66-77.9) |
| With XBB.1.5 vaccination | 48.8 (36.4-58.8) | 67.7 (61.2-73.1) | 85.3 (80.6-88.9) |

^a^ Adjusted for age group (18-39, 40-59, 60-69, 70-85 years), sex, education level and medical risk condition

**Figure S2**. Flowchart of total number of infections and sequenced self-tests in the period 9 October 2023 to 9 January 2024

Total number of VASCO participants in the analysis: 23,895

Positive test reported: 3,525 (14.8%)

ICF provided for sequencing: 2,560 (72.6%)

Self-tests send in for sequencing: 846 (33.0%)

Included in sequencinga: 764 (90.3%)

(near) whole genome sequencing: 530 (69.4%)

^a^ Weak-positive self-tests were not included in sequencing

**Figure S3**. Variants of infection based on sequencing of self-tests per week from 9 October 2023 to 9 January 2024

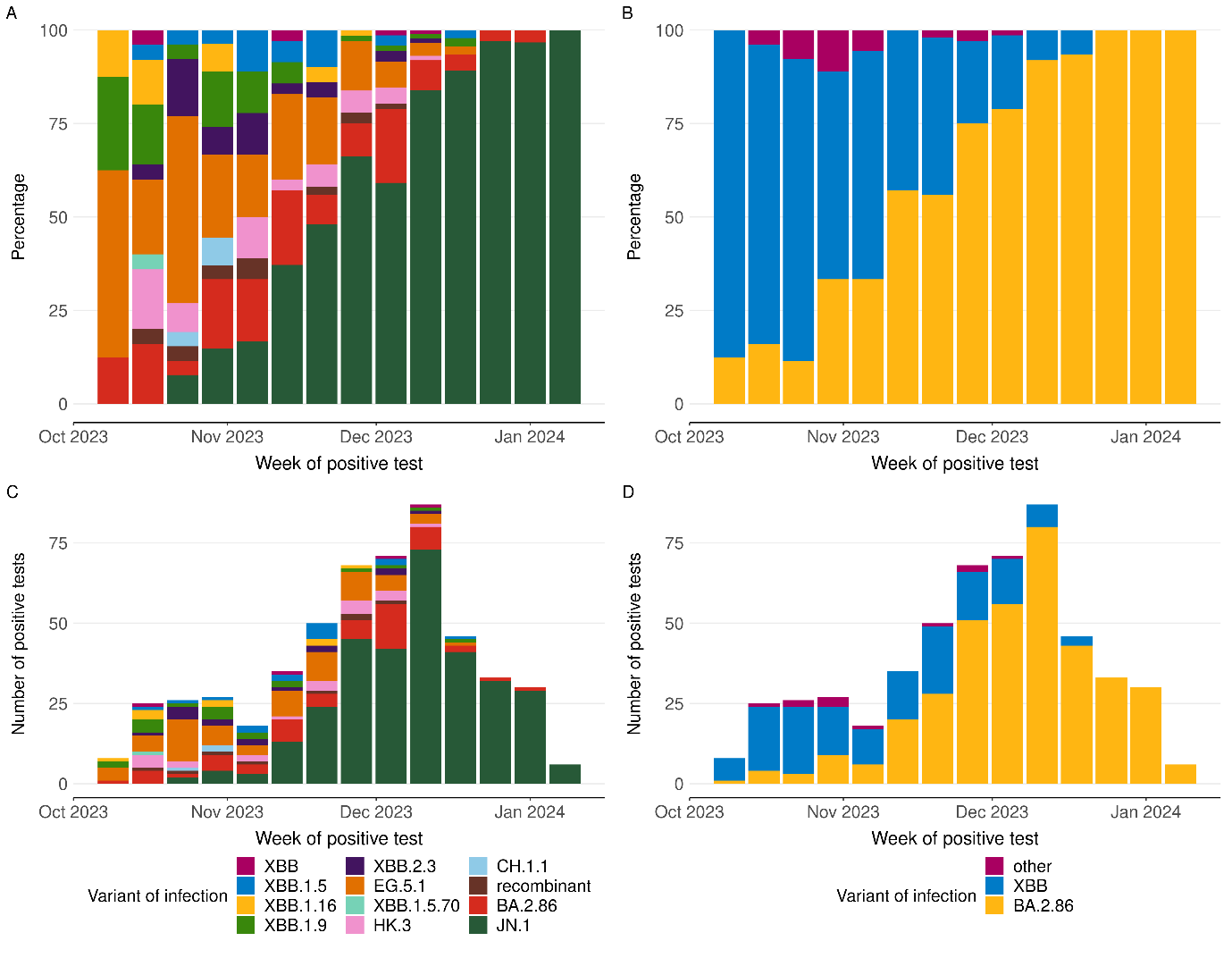

A) and C) SARS-COV-2 subvariants of infection observed during the study period; B) and D) general classification of subvariants of infection observed during the study period, with all XBB subvariants (XBB, XBB.1.5, XBB.1.16, XBB.1.9, XBB.2.3, EG.5.1, XBB.1.5.70, HK.3) and all BA.2.86 derivatives (BA.2.86, JN.1, JN.2, JN.3, JN.4, JN.5, JN.6, JN.9, JN.10 and subvariants) grouped together for the XBB vs BA.2.86 analysis.

**Table S3**. Demographics of participants by variant of infection based on sequencing of self-tests in the period 9 October 2023 to 9 January 2024

|  | **Total** | | **BA.2.86 infection** | | **XBB infection** | | **P-value** |
| --- | --- | --- | --- | --- | --- | --- | --- |
|  | n | % | n | % | n | % |  |
| **All participants** | 519 | 100 | 370 | 100 | 149 | 100 |  |
| **Age (years)** | | | | | | | |
| 18-59 years | 173 | 33.3 | 114 | 30.8 | 59 | 39.6 | 0.069 |
| 60-85 years | 346 | 66.7 | 256 | 69.2 | 90 | 60.4 |  |
| **Gender** | | | | | | | |
| Female | 370 | 71.3 | 267 | 72.2 | 103 | 69.1 | 0.559 |
| Male | 149 | 28.7 | 103 | 27.8 | 46 | 30.9 |  |
| **Medical risk condition, yes** | 215 | 41.4 | 158 | 42.7 | 57 | 38.3 | 0.405 |
| **Education level** | | | | | | | |
| High | 309 | 59.5 | 217 | 58.6 | 92 | 61.7 | 0.587 |
| Intermediate | 146 | 28.1 | 105 | 28.4 | 41 | 27.5 |  |
| Low | 60 | 11.6 | 46 | 12.4 | 14 | 9.4 |  |
| Other | 4 | 0.8 | 2 | 0.5 | 2 | 1.3 |  |
| **XBB.1.5 vaccination** | | | | | | | |
| Yes | 228 | 43.9 | 197 | 53.2 | 31 | 20.8 | <0.001 |
| No | 291 | 56.1 | 173 | 46.8 | 118 | 79.2 |  |
| **Infection history** |  | |  | |  | |  |
| No prior infection | 100 | 19.3 | 65 | 17.6 | 35 | 23.5 | 0.019 |
| Infection > 1 year ago | 327 | 63.0 | 229 | 61.9 | 98 | 65.8 |  |
| Infection < 1 year ago | 92 | 17.7 | 76 | 20.5 | 16 | 10.7 |  |
